# The association of Red Cell Distribution Width and Red Cell Distribution Width related indices with the in-Hospital Mortality of Congestive Heart Failure in a retrospective observational cohort study

**DOI:** 10.64898/2026.05.29.26354291

**Authors:** Dongxia Wang, Dingchao Lv, Xin Yuan, Yuyuan Wang

**Affiliations:** Department of Cardiology, Shanxi Provincial People’s Hospital, The Affiliated Hospital of Shanxi Medical University, Taiyuan, Shanxi, China; Department of Cardiology, Changzhi People’s Hospital, The Affiliated Hospital of Changzhi Medical College, Changzhi, Shanxi, China; Department of Critical Care Medicine, Changzhi People’s Hospital, The Affiliated Hospital of Changzhi Medical College, Changzhi, Shanxi, China

**Keywords:** in-hospital mortality, congestive heart failure, RDW, RDW related indices, Positive association

## Abstract

**Background:** Red cell distribution width (RDW), a readily available hematological parameter reflecting erythrocyte size heterogeneity, has been increasingly recognized as a prognostic marker in congestive heart failure (CHF), with elevated levels independently associated with adverse outcomes. However, RDW-derived composite indices—particularly the RDW-to-platelet ratio (RPR) and RDW-to-hemoglobin ratio (RHR), which integrate inflammatory, hemostatic, and oxygen-delivery pathways—remain largely unexplored in CHF populations. Whether these indices provide incremental prognostic value beyond RDW alone in critically ill patients with CHF has not been established.

**Methods:** This retrospective cohort study included 30,409 participants from the MIMIC-IV and eICUCRD databases. Multivariable logistic regression, restricted cubic spline (RCS) analysis, and subgroup analyses were employed to evaluate the associations between RDW, RDWderived indices (RPR and RHR), and in-hospital mortality in patients with congestive heart failure.

**Results:** Based on a pooled cohort of 30,409 patients with CHF from the MIMIC-IV and multi-center eICU-CRD databases (15,983 and 14,426, respectively), 16,295 (53.6%) were male and 14,114 were female, with a median age of 71.7 years. The mean RDW was 16.0 ± 2.5, and the overall in-hospital mortality rate was 12.6%. Higher RDW quintiles were associated with progressively increased in-hospital mortality. In the fully adjusted model, RDW, RPR, and RHR were all significantly associated with increased in-hospital mortality, with adjusted odds ratios (ORs) of 2.46 (95% CI: 2.17–2.79) for RDW,1.55 (95% CI: 1.38–1.73) for RPR, and 2.43 (95% CI: 2.09–2.82) for RHR. Sensitivity analyses using restricted cubic splines demonstrated that the association between RDW and RHR with in-hospital mortality was linear (P for nonlinearity > 0.05), whereas that for RPR exhibited a non-linear pattern (P = 0.02 for nonlinearity).

**Conclusions:** Elevated RDW, RPR, and RHR were independently associated with increased in-hospital mortality in patients with congestive heart failure. Notably, RPR exhibited a non-linear threshold association with in-hospital mortality.

## Introduction

### Background

Congestive heart failure (CHF) represents one of the most formidable challenges in contemporary cardiovascular medicine, with a global prevalence exceeding 64 million cases—a figure anticipated to climb steeply as populations age and the epidemiological burden of hypertension, coronary artery disease, and diabetes intensifies[1]. Although landmark pharmacological advances— namely angiotensin receptor-neprilysin inhibitors, sodium-glucose cotransporter 2 inhibitors, and mineralocorticoid receptor antagonists— have reshaped chronic disease management, outcomes remain discouraging: 5-year mortality still surpasses 50%, and nearly one in four patients is rehospitalized within 30 days[1, 2]. Current risk stratification hinges largely on clinical prediction models such as the MAGGIC and Seattle Heart Failure Model[3, 4], often supplemented by natriuretic peptides or high-sensitivity troponin; yet these instruments were derived predominantly from ambulatory cohorts and reflect single biological axes, limiting their transferability to the critically ill. In the ICU, where acute decompensation, multiorgan failure, and hemodynamic instability intersect simultaneously, there is an unmet need for inexpensive, rapidly available biomarkers that capture the multidimensional pathophysiology driving mortality.

Red cell distribution width (RDW), a standard component of the complete blood count that quantifies erythrocyte size variability, has garnered increasing attention as a widely available and inexpensive prognostic indicator across a broad spectrum of cardiovascular and non-cardiovascular conditions[5]. The biological rationale for this association lies in the fact that RDW mirrors interconnected pathophysiological processes—including impaired erythropoiesis, iron deficiency, chronic inflammation, and oxidative stress—that converge to accelerate adverse cardiovascular events[6]. Within HF cohorts, multiple large-scale observational analyses have confirmed RDW as an independent predictor of all-cause mortality, yielding adjusted hazard ratios between 1.11 and 1.29, and have shown that adding RDW to models containing N-terminal pro-brain natriuretic peptide or left ventricular ejection fraction improves risk discrimination[7]. Acknowledging that any single parameter captures only a narrow biological window, investigators have increasingly turned to composite indices that pair RDW with complementary hematological variables. The RDW-to-platelet ratio (RPR), derived by dividing RDW (fL) by platelet count (×10^9^/L), reflects the joint burden of erythrocyte dysregulation and thrombotic predisposition. Among patients with ST-segment elevation myocardial infarction treated by primary percutaneous coronary intervention, an RPR exceeding 0.061 independently predicted oneyear cardiovascular mortality (OR = 3.106, 95% CI: 1.456–6.623) and major adverse cardiovascular events[8]. In parallel, a lower hemoglobin-to-RDW ratio (HRR) has been linked to heightened in-hospital mortality risk in chronic HF populations[9]. Beyond platelet- and hemoglobin-based constructs, the RDW-to-albumin ratio (RAR) captures both inflammatory and nutritional dimensions; analyses of the MIMIC-III and MIMIC-IV intensive care databases demonstrated that RAR surpassed either component alone in predicting inhospital mortality (AUC = 0.683 and 0.710, respectively; optimal cut-off 4.5–5.0), reinforcing its potential as a pragmatic bedside risk stratification tool[10]. Taken together, these RDWbased composite indices—calculated from universally available complete blood count and biochemical parameters—offer a promising approach to refining cardiovascular risk prediction, yet they require further validation in heterogeneous clinical settings.

### Objectives

Despite the biological appeal of RDW-derived composite indices, both the RPR and the RDW-to-hemoglobin ratio (RHR)—which jointly capture inflammatory, hemostatic, and oxygen-delivery dynamics—remain conspicuously understudied in CHF populations, leaving a pronounced gap in the existing literature. Accordingly, this study was designed to examine the associations of RDW and its two derived ratios (RPR and RHR) with in-hospital mortality among patients with severe CHF. Methods:

### Data Sources

This study adopted a retrospective observational design using a large, publicly available repository. The dataset (doi: 10.5061/dryad.tx95x6b18) comprised data from two distinct multi-center critical care databases: the Medical Information Mart for Intensive Care IV (MIMIC-IV), which contains more than 40,000 ICU admissions from Beth Israel Deaconess Medical Center in Boston between 2008 and 2019[11], and the eICU Collaborative Research Database (eICU-CRD), which incorporates de-identified records from over 200,000 ICU admissions across 208 hospitals in the United States between 2014 and 2015[12]. Both repositories provide extensive clinical documentation encompassing demographic variables, high-resolution physiological measurements, ICD-9-coded diagnoses, and laboratory findings[12]. All records were de-identified in accordance with the Health Insurance Portability and Accountability Act (HIPAA) Safe Harbor provisions, and the requirement for informed consent was waived. The study followed the Strengthening the Reporting of Observational Studies in Epidemiology (STROBE) guidelines.

### Study population

Initially, 30,474 patients with CHF undergoing their first ICU admission were identified via ICD-9 code 4280 (MIMIC-IV: n = 16,012; eICU: n = 14,462). Patients were subsequently excluded if they were younger than 18 years, had missing hemoglobin, platelet, or RDW values, or lacked outcome information. After these exclusions, 30,409 patients constituted the final analytic cohort (MIMIC-IV: n = 15,983; eICU: n = 14,426) (Supplementary Materials Figure 1).

### Data Extraction

Data extraction procedures have been detailed elsewhere[11]. Within the eICU database, 2 patients younger than 18 years were excluded. The following variables were extracted: (1) demographic characteristics—age, ethnicity, and sex; (2) vital signs recorded within 24 hours of ICU admission—systolic blood pressure (SBP), mean blood pressure (MBP), respiratory rate (RR), heart rate (HR), and body temperature; (3) comorbid conditions—chronic obstructive pulmonary disease (COPD), diabetes mellitus, hepatic failure (HepF), and acute myocardial infarction (AMI); (4) laboratory values obtained within 24 hours of ICU admission—anion gap (AG), blood urea nitrogen (BUN), calcium, chloride, creatinine, potassium, sodium, hemoglobin (Hb), mean corpuscular hemoglobin (MCH), mean corpuscular hemoglobin concentration (MCHC), mean corpuscular volume (MCV), platelet count, red blood cell (RBC) count, red cell distribution width (RDW), and white blood cell (WBC) count; (5) severity indices—Sequential Organ Failure Assessment (SOFA) and Acute Physiology Score III (APS III); (6) vasoactive agents—norepinephrine, dopamine, epinephrine, phenylephrine, and vasopressin; (7) respiratory support—mechanical ventilation and intubation; and (8) derived ratios— RHR (RDW [%] ÷ Hb [g/L]) and RPR (RDW [%] ÷ platelet count [×10^9^/L])[8].

### Outcome

In-hospital mortality, defined as death from any cause prior to discharge from the index hospitalization, constituted the primary outcome.

### Statistical analysis

Normally distributed continuous data are reported as mean ± SD, whereas values deviating from normality are summarized by median (IQR). Categorical variables appear as frequencies (percentages). Depending on the outcome of the Shapiro–Wilk test, group comparisons for continuous variables used either the independent-samples t-test or the Mann–Whitney U test. For categorical variables, the chi-square test or Fisher’s exact test was applied, as warranted.

To examine the independent relations of RDW and its derived indices with in-hospital mortality, univariable followed by multivariable logistic regression was performed. Candidate covariates were screened on the basis of clinical relevance and univariable significance (p < 0.05), while respecting biologically plausible mechanistic pathways. Three sequentially adjusted multivariable logistic regression models were then constructed: Model 1 adjusted for age, sex, and race; Model 2 additionally incorporated COPD, diabetes mellitus, hepatic failure, heart rate, and systolic blood pressure; and Model 3 further adjusted for anion gap, blood urea nitrogen, calcium, potassium, red blood cell count, sodium, and white blood cell count. Predefined subgroup analyses were undertaken to appraise the consistency of the observed associations, thereby corroborating the robustness of the primary findings.

Restricted cubic spline (RCS) regression was applied to explore potential nonlinear dose-response relations between the indices and in-house mortality. Whenever nonlinearity was evident, piecewise regression was implemented to pinpoint the inflection point and characterize the association.

Subgroup and interaction analyses were conducted according to sex, age (<65 vs. ≥65 years), diabetes, and COPD to explore potential effect modification. All subgroup models incorporated adjustment for age, sex, race, heart rate, systolic blood pressure, anion gap, blood urea nitrogen, calcium, potassium, red blood cell count, sodium, white blood cell count, diabetes mellitus, COPD, and hepatic failure. Findings are illustrated as odds ratios (ORs) alongside 95% confidence intervals (CIs) in forest plots.

## Results

### Baseline characteristic of participants

The analytic cohort comprised 30,409 patients with CHF (MIMIC-IV: 15,983 [52.6%]; eICU: 14,426 [47.4%]), with a mean age of 71.7 ± 13.7 years and a male predominance of 16,295 (53.6%). The cohort-wide mean RDW was 16.0 ± 2.5%, and the overall in-hospital mortality rate reached 12.6%. Individuals in higher RDW strata exhibited a clinical profile that diverged markedly from their lower-RDW counterparts. Notably, they were more likely to receive vasoactive agents yet less frequently underwent mechanical ventilation or intubation.

Comorbidity burden was comparatively lighter in this subgroup. Severity-of-illness assessments revealed higher SOFA and APS III scores, accelerated heart rates, and diminished blood pressures among patients with elevated RDW. From a laboratory standpoint, higher RDW correlated with reduced chloride, hemoglobin, MCH, MCHC, MCV, and WBC, coupled with elevated BUN, creatinine, and RDW itself.

Non-survivors displayed markedly greater levels of RDW, RPR, and RHR relative to survivors (Figure 1), with mean values of 16.90 versus 15.88 for RDW, 0.13 versus 0.10 for RPR, and 1.72 versus 1.58 for RHR.

**Fig. 1.**
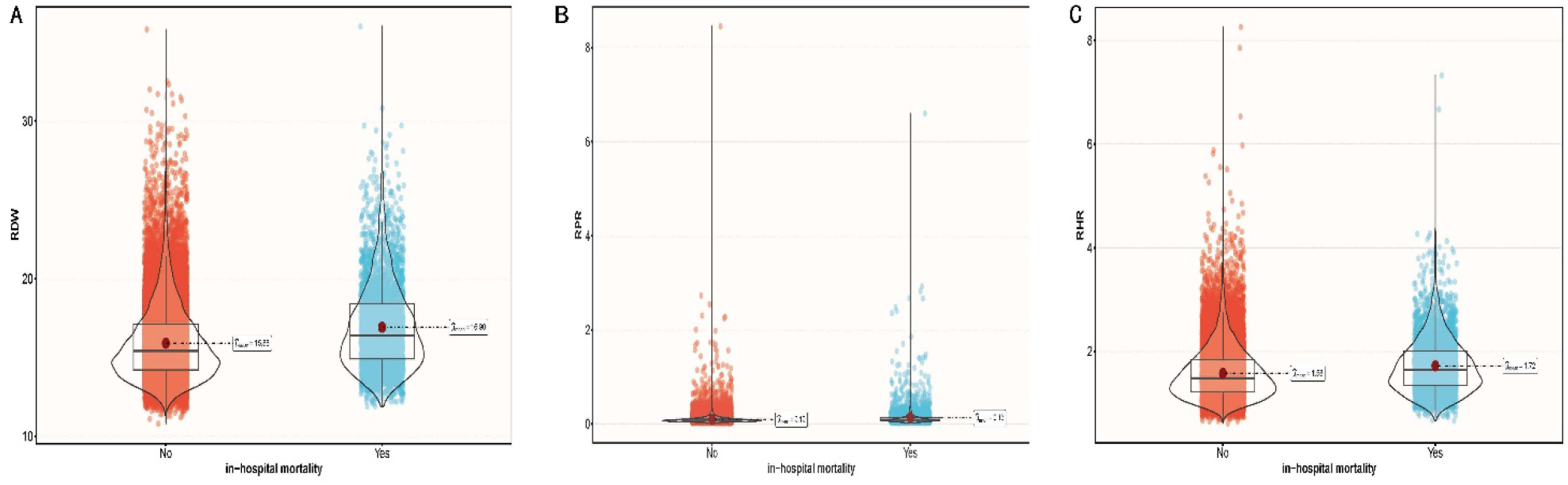
Difference in distribution of RDW (A), RPR(B), RHR(C) in alive and expired groups. The solid gray line in the middle of the Box Plot represents the median, and the upper and lower edge lines of the box plot represent the first quartile and the third quartile. The confluence of the two ends of the Violin Plot represents the lower adjusted value and upper adjacent value. The linear parts of the Violin Plot at the upper and lower ends reflect the outside points. The scatter plots in the red and blue sections visually reflect the distribution of data. RPR: RDW and platelets ratio, RHR: RDW and Hb ratio.

The association of RDW and RDW related indices with in-hospital mortality in all participants

After comprehensive covariate adjustment (Model 3, Table 2), each of RDW, RPR, and RHR emerged as an independent predictor of in-hospital mortality, with corresponding odds ratios of 1.12 (95% CI: 1.11–1.14; P < 0.001) for RDW, 1.18 (95% CI: 1.15–1.21; P < 0.001) for RPR, and 1.58 (95% CI: 1.45–1.71; P < 0.001) for RHR. For clinical interpretability, RPR was rescaled by a factor of 10, such that the OR reflects the change in odds associated with each 0.1-unit increment in the original value.

**Table 1.**
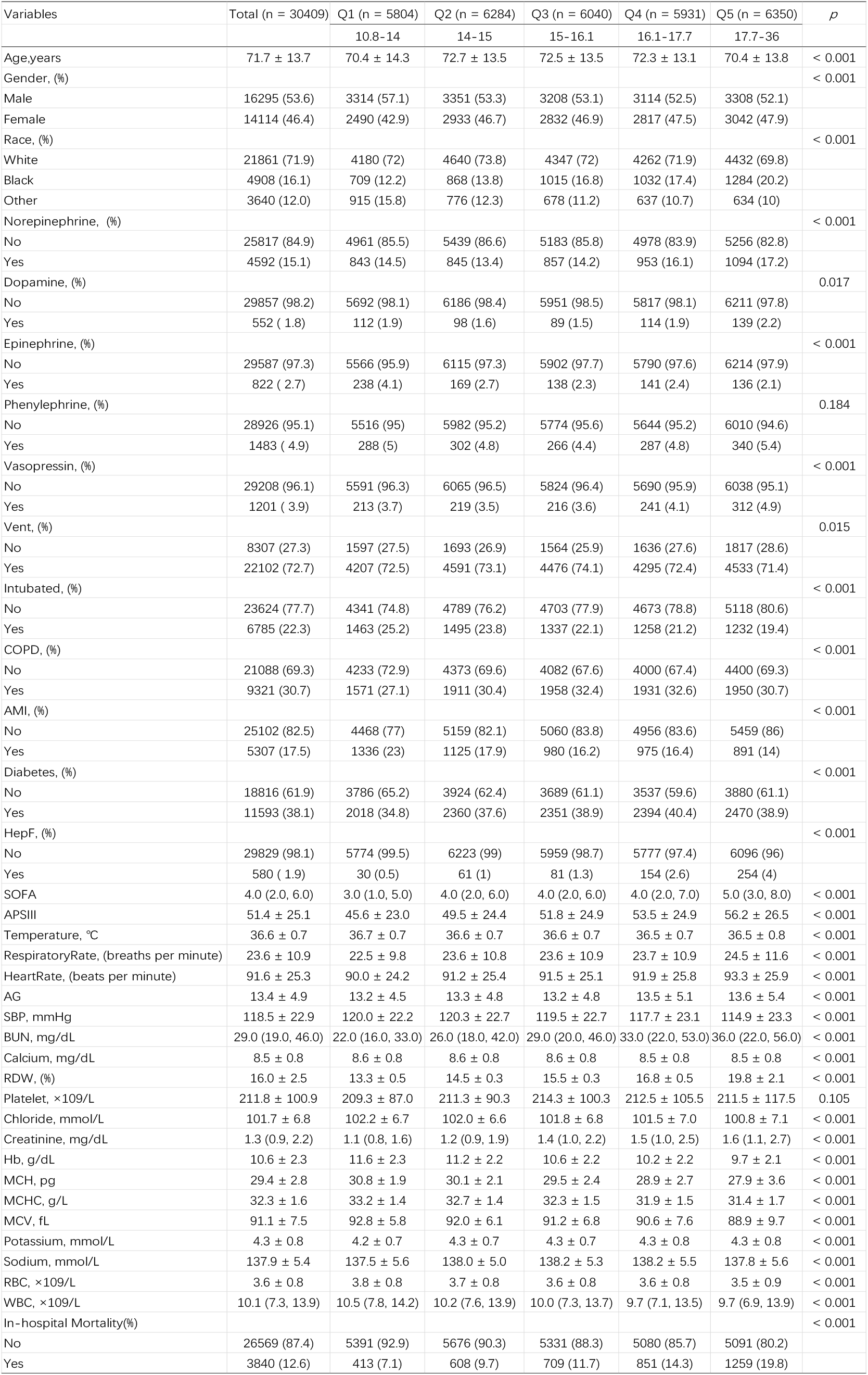
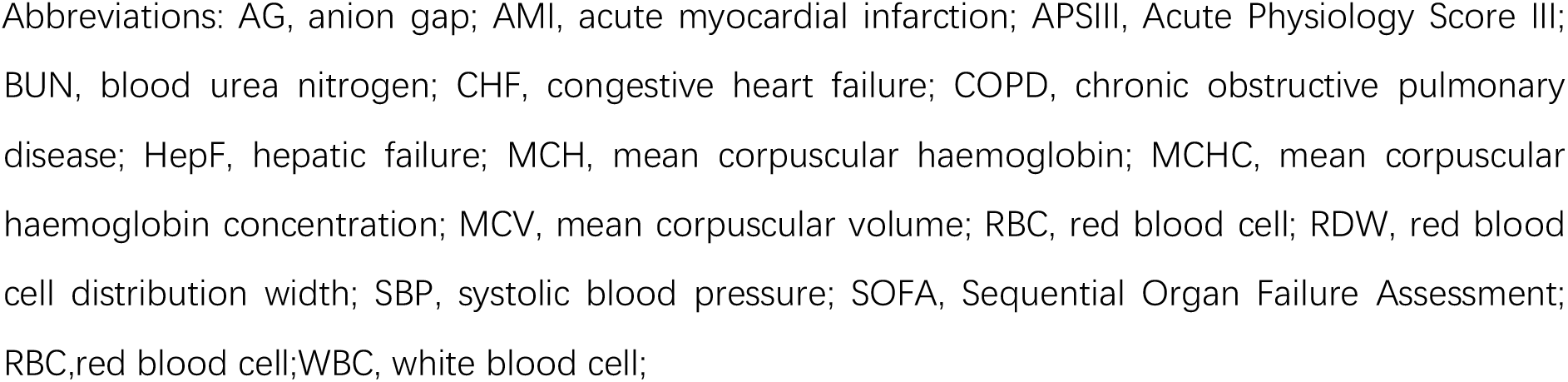
Baseline characteristics of included subjects stratified by baseline RDW index.

**Table 2.** The association of RDW, RPR*10, and RHR with in-hospital mortality in all participants.

| Variable | Crude |  | Model1 |  | Model2 |  | Model3 |  |
| --- | --- | --- | --- | --- | --- | --- | --- | --- |
|  | OR (95%CI) | Pvalue | OR (95%CI) | Pvalue | OR (95%CI) | Pvalue | OR (95%CI) | Pvalue |
| RDW | 1.15 (1.14~1.17) | <0.001 | 1.17 (1.15~1.18) | <0.001 | 1.14 (1.13~1.16) | <0.001 | 1.12 (1.11~1.14) | <0.001 |
| Quintile |  |  |  |  |  |  |  |  |
| Q1 | 1 (Ref) | <0.001 | 1 (Ref) |  | 1 (Ref) |  | 1 (Ref) |  |
| Q2 | 1.4 (1.23~1.59) | <0.001 | 1.34 (1.18~1.53) | <0.001 | 1.31 (1.15~1.5) | <0.001 | 1.23 (1.08~1.41) | 0.002 |
| Q3 | 1.74 (1.53~1.97) | <0.001 | 1.7 (1.49~1.93) | <0.001 | 1.62 (1.42~1.84) | <0.001 | 1.48 (1.3~1.69) | <0.001 |
| Q4 | 2.19 (1.93~2.47) | <0.001 | 2.16 (1.91~2.45) | <0.001 | 1.99 (1.76~2.26) | <0.001 | 1.7 (1.49~1.94) | <0.001 |
| Q5 | 3.23 (2.87~3.63) | <0.001 | 3.39 (3.01~3.82) | <0.001 | 2.95 (2.62~3.33) | <0.001 | 2.46 (2.17~2.79) | <0.001 |
| P for trend |  | <0.001 |  | <0.001 |  | <0.001 |  | <0.001 |
| RPR*10 | 1.22 (1.18~1.25) | <0.001 | 1.23 (1.19~1.26) | <0.001 | 1.19 (1.16~1.23) | <0.001 | 1.18 (1.15~1.21) | <0.001 |
| Quintile |  |  |  |  |  |  |  |  |
| Q1 | 1 (Ref) |  | 1 (Ref) |  | 1 (Ref) |  | 1 (Ref) |  |
| Q2 | 0.82 (0.73~0.92) | 0.001 | 0.8 (0.72~0.9) | <0.001 | 0.83 (0.74~0.93) | 0.002 | 0.9 (0.79~1.01) | 0.076 |
| Q3 | 0.95 (0.85~1.06) | 0.35 | 0.92 (0.82~1.03) | 0.128 | 0.93 (0.83~1.05) | 0.244 | 1 (0.89~1.13) | 0.943 |
| Q4 | 1.05 (0.94~1.17) | 0.391 | 1 (0.89~1.11) | 0.951 | 0.99 (0.89~1.11) | 0.916 | 1.07 (0.95~1.2) | 0.252 |
| Q5 | 1.7 (1.54~1.89) | <0.001 | 1.65 (1.48~1.82) | <0.001 | 1.52 (1.37~1.69) | <0.001 | 1.55 (1.38~1.73) | <0.001 |
| P for trend |  | <0.001 |  | <0.001 |  | <0.001 |  | <0.001 |
| RHR | 1.64 (1.55~1.75) | <0.001 | 1.66 (1.56~1.76) | <0.001 | 1.6 (1.5~1.7) | <0.001 | 1.58 (1.45~1.71) | <0.001 |
| Quintile |  |  |  |  |  |  |  |  |
| Q1 | 1 (Ref) |  | 1 (Ref) |  | 1 (Ref) |  | 1 (Ref) |  |
| Q2 | 1.28 (1.13~1.45) | <0.001 | 1.21 (1.07~1.37) | 0.002 | 1.24 (1.1~1.41) | 0.001 | 1.32 (1.16~1.51) | <0.001 |
| Q3 | 1.53 (1.36~1.72) | <0.001 | 1.44 (1.28~1.63) | <0.001 | 1.51 (1.33~1.7) | <0.001 | 1.61 (1.41~1.85) | <0.001 |
| Q4 | 1.82 (1.62~2.04) | <0.001 | 1.73 (1.54~1.95) | <0.001 | 1.75 (1.55~1.97) | <0.001 | 1.86 (1.61~2.13) | <0.001 |
| Q5 | 2.35 (2.1~2.63) | <0.001 | 2.3 (2.05~2.57) | <0.001 | 2.25 (2.01~2.53) | <0.001 | 2.43 (2.09~2.82) | <0.001 |
| P for trend |  | <0.001 |  | <0.001 |  | <0.001 |  | <0.001 |

Crude model: adjusted for none;
Model 1 was adjusted only for age, gender, and race;
Model 2 was additionally adjusted for COPD, diabetes, liver failure, HR, and SBP;
Model 3 was additionally adjusted for AG, BUN, calcium, potassium, RBC, sodium, and WBC.

**Table 3.** The non-linearity relationship between RPR*10 and the risk of in-hospital mortality.

| Threshold of RPR*10 | OR | 95%CI | P-value |
| --- | --- | --- | --- |
| <0.524 | 1.134 | 0.341, 3.772 | 0.8381 |
| ≥0.524 | 1.425 | 1.301, 1.562 | <0.001 |

Within the fully adjusted model (Model 3, Table 2), patients in the uppermost quintile of each index carried substantially greater odds of in-hospital mortality than those in the bottom quintile, yielding adjusted ORs of 2.46 (95% CI: 2.17–2.79) for RDW, 1.55 (95% CI: 1.38–1.73) for RPR, and 2.43 (95% CI: 2.09–2.82) for RHR.

RCS curves (Figure 2) revealed divergent dose-response patterns: RPR exhibited a significant nonlinear relation with in-hospital mortality (P for nonlinearity < 0.05), whereas RDW and RHR approximated linearity (P for nonlinearity > 0.05), all within the fully adjusted model (Model 3, Table 2). A two-piecewise regression was consequently fitted for RPR, identifying an inflection point at 0.524 (Table 5). Beyond this threshold, each unit rise in RPR conferred a 42.5% increase in the odds of death (OR = 1.425; 95% CI: 1.301–1.562), whereas below 0.524 the association was attenuated and no longer significant (OR = 1.134; 95% CI: 0.341–3.772).

**Fig. 2.**
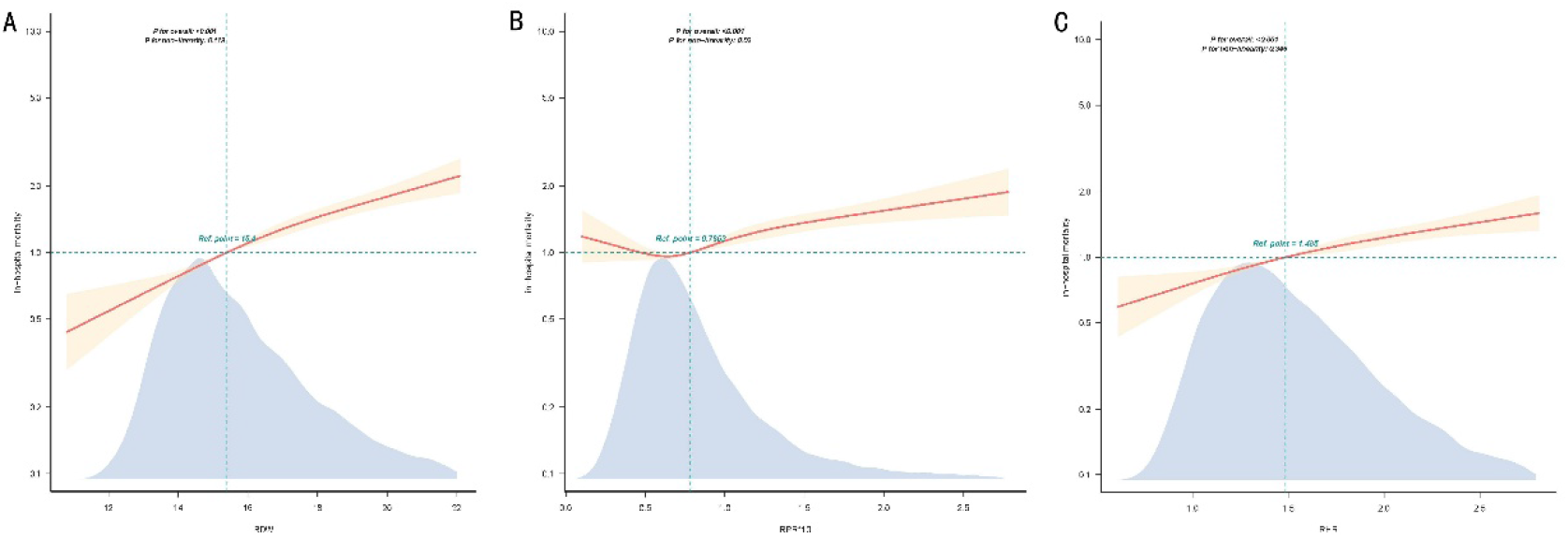
Restricted cubic spline curves for heart disease by RDW (A), RPR*10 (B), RHR(C) in all participants after covariate adjustment. Heavy central line represents the estimated adjusted odd ratio, with shaded ribbons denoting 95% confidence interval. The model is adjusted for age, gender, race,COPD, diabetes, liver failure, HR, SBP,AG, BUN, calcium, potassium, RBC, sodium, and WBC.

### Subgroup analysis

Subgroup analyses explored whether the prognostic impact of RDW, RPR, and RHR varied across prespecified strata (Figure 3A–C), encompassing age (<65 vs. ≥65 years), sex, race, diabetes mellitus, COPD, and AMI. Interaction terms were assessed via likelihood ratio testing. A significant age-by-RDW interaction emerged (P < 0.05), suggesting differential mortality risk across age strata; all remaining subgroup interactions lacked statistical significance (sex, race, diabetes mellitus, COPD, and AMI; all P > 0.05).

**Fig. 3.**
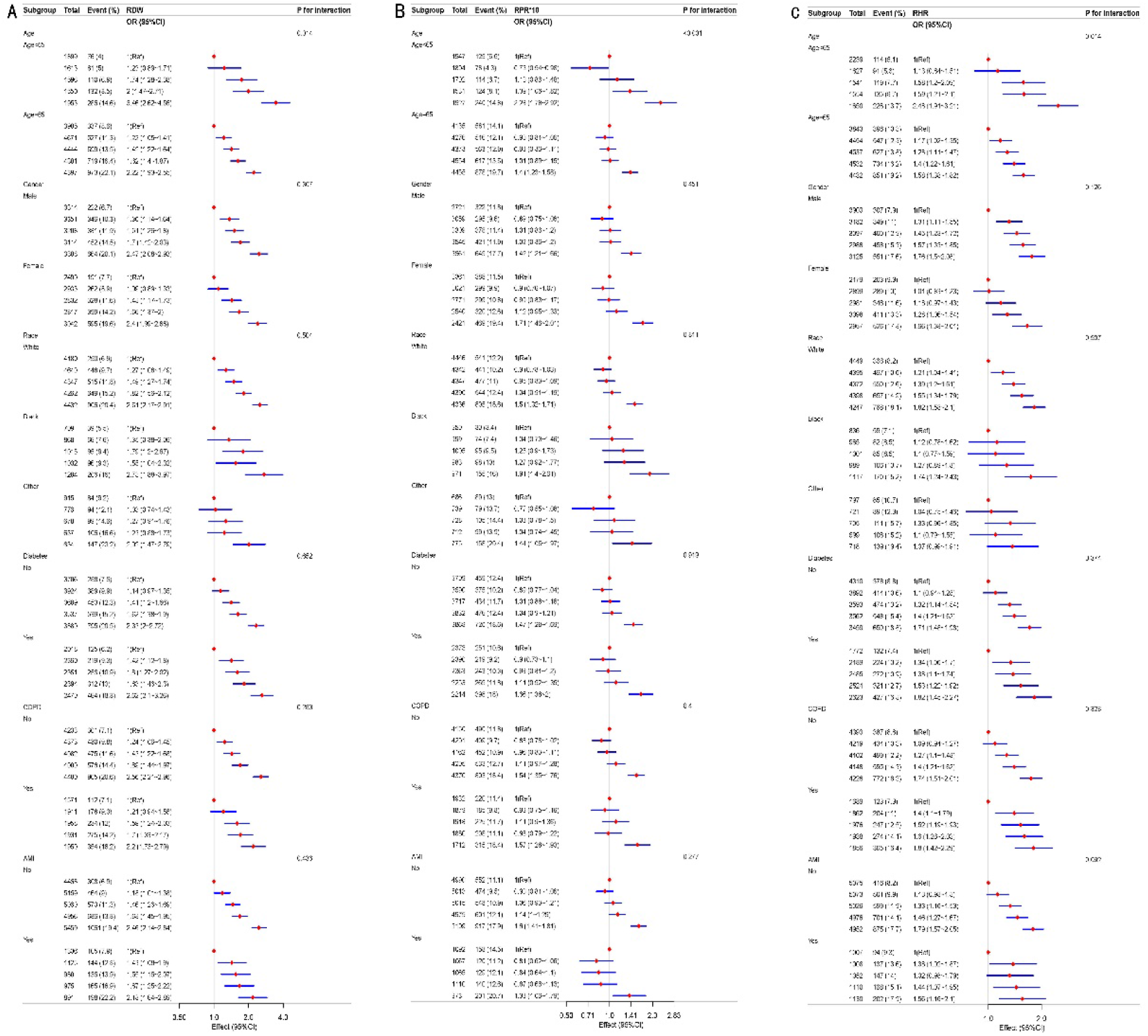
Subgroup analyses in the chronic heart failure population for the association of RDW (A), RPR*10 (B) and RHR (C) with risk of in-hospital mortality in age, gender, race, diabetes, COPD, and AMI subgroups. The OR was calculated using Logistic proportional hazards model with the adjustments including age, gender, race,COPD, diabetes, liver failure, HR, SBP,AG, BUN, calcium, potassium, RBC, sodium and WBC. The P values for the interaction effect that using the likelihood ratio test are annotated to the right of A, B, and C. OR odds ratio, CI confidence interval

## Discussion

CHF imposes a formidable and steadily mounting global health burden, affecting in excess of 64 million individuals worldwide. Its prevalence continues to climb, propelled principally by population aging and enhanced post-acute cardiovascular event survival[13]. Notwithstanding marked progress in guideline-directed medical therapy, CHF prognosis remains dismal, with a 5-year mortality rate surpassing 50%[14]. Prevailing risk stratification hinges largely on clinical scoring systems—such as the MAGGIC and Seattle Heart Failure Models—augmented by established biomarkers including B-type natriuretic peptide (BNP) and NT-proBNP[15]. These instruments, however, exhibit constrained sensitivity and specificity for individual outcome prediction, particularly among patients with multimorbidity[16]. Furthermore, conventional prognostic models may incompletely reflect the broader pathophysiological landscape underpinning disease progression, encompassing systemic inflammation, nutritional compromise, and renal dysfunction[5]. Accordingly, a compelling unmet need persists for novel, readily accessible biomarkers that can refine risk stratification and steer personalized management strategies in CHF.

Red cell distribution width (RDW) is a standard hematological index that quantifies anisocytosis (erythrocytic size variation). Although RDW has conventionally been employed in the differential diagnosis of anemia, mounting evidence now positions it as an independent prognostic marker across an array of cardiovascular diseases, including CHF[5]. Among patients with CHF, a higher RDW mirrors underlying pathophysiological perturbations—encompassing chronic inflammation, oxidative stress, neurohormonal activation, and impaired bone marrow erythropoiesis—each of which propels adverse cardiac remodeling and presages poorer clinical outcomes[7].

Platelet abnormalities are likewise frequent in CHF, wherein endothelial dysfunction and low-grade inflammation drive platelet activation and perturbed kinetics, engendering a prothrombotic milieu that amplifies cardiovascular risk[17]. In parallel, anemia—irrespective of whether it stems from iron deficiency, renal dysfunction, or hemodilution—afflicts a sizeable fraction of CHF patients and portends heightened morbidity and mortality[18]. Since RDW, platelet indices, and hemoglobin each interrogate discrete yet converging pathways within CHF pathophysiology, integrative composite indices may yield superior prognostic discrimination compared with any solitary marker[8].

Within this multicenter cohort, RDW, RHR, and RPR were each linked to heightened inhospital mortality risk among patients with CHF, and these associations persisted after full covariate adjustment.Several mechanistic pathways may underlie the observed relations between elevated RDW-derived indices and in-hospital death in CHF. An elevated RPR embodies a dual pathological process: accentuated erythrocyte heterogeneity—reflecting chronic inflammation, oxidative stress, and impaired erythropoiesis—paired with thrombocytopenia that may denote bone marrow suppression, consumptive coagulopathy, or immune-mediated platelet destruction[6]. Within the CHF milieu, a high RPR therefore heralds a compounded inflammatory–hemostatic deficit: patients concurrently shoulder a heavier inflammatory burden and a diminished hemostatic reserve, rendering them more susceptible to acute decompensation, bleeding complications, and organ hypoperfusion. Importantly, notwithstanding reduced platelet counts, platelet activation—rather than platelet number per se—orchestrates the prothrombotic state in heart failure, forging a paradoxical landscape wherein patients contend with both thrombotic and hemorrhagic risks[19].

Likewise, an elevated RHR unites two interrelated prognostic signals. A higher RDW mirrors systemic inflammatory and oxidative stress that perturbs erythropoiesis[6], whereas reduced hemoglobin—whether consequent to genuine iron deficiency, renal insufficiency, or hemodilution— compromises tissue oxygen delivery[20]. Within CHF, anemia intensifies the already precarious equilibrium between myocardial oxygen supply and demand, augmenting cardiac workload and provoking progressive decompensation. The concurrence of a high RDW and low hemoglobin consequently pinpoints patients burdened by both amplified inflammatory stress and impaired oxygen-carrying capacity—a particularly lethal tandem in the crucible of critical illness. Importantly, this dual-pathway integration parallels the established prognostic merit of composite RDW–platelet indices in acute coronary syndromes[8], lending credence to the utility of combined hematological ratios in risk stratification.

Importantly, the present analysis uncovered a U-shaped relation between RPR and inhospital mortality in CHF—a pattern hitherto unreported in the literature. At the lower end of the RPR spectrum, diminished RDW coupled with preserved platelet counts may denote a comparatively stable hematological phenotype; nevertheless, exceedingly low RPR values could equally signal reactive thrombocytosis secondary to acute inflammation, which paradoxically elevates cardiovascular risk[21, 22]. By contrast, at the higher end of the RPR spectrum, elevated RDW coexisting with thrombocytopenia portends compounded inflammatory–hemostatic dysfunction[19], as delineated earlier. This biphasic signature intimates that both RPR extremes may impart adverse prognostic implications through divergent mechanisms—reactive thrombosis at the low extreme versus hemostatic failure at the high extreme—underscoring the imperative of appraising the entire RPR continuum rather than a unidirectional threshold in clinical risk assessment.

The significant age-by-RDW interaction detected in our analysis may stem from several interrelated mechanisms. First, RDW is documented to climb with advancing age, owing to age-related bone marrow attrition, chronic low-grade inflammation (dubbed “inflammaging”), and a higher prevalence of subclinical iron deficiency—all of which may blunt its discriminative capacity for mortality prediction in older patients[23]. Second, younger CHF patients more commonly harbor ischemic or genetic etiologies characterized by a more uniform pathophysiology, whereas older patients display greater heterogeneity, encompassing frailty, multiorgan dysfunction, and non-cardiac drivers of mortality, potentially magnifying RDW’s prognostic resonance[6, 24]. Third, platelet function and coagulation cascades experience marked age-related alterations, which may modulate the inflammatory-thrombotic crosstalk that anchors RDW’s association with outcomes. Taken together, these observations highlight that RDW’s prognostic utility may be age-contingent, warranting age-stratified risk stratification in ensuing studies.

Several constraints inherent to this study merit delineation. First, the adoption of a retrospective observational architecture precludes causal inference. Second, RDW, platelet count, and hemoglobin concentrations were captured solely as baseline measurements within the inaugural 24-hour window following ICU admission, a methodology that may fail to encapsulate dynamic physiological trajectories during the hospital course. Finally, pivotal echocardiographic metrics—LVEF foremost among them—remained inaccessible within the eICU-CRD, thereby foreclosing the possibility of stratifying analyses according to heart failure subtype or severity.

## Conclusions

Within this multicenter cohort, elevated RDW, RPR, and RHR were each linked to heightened in-hospital mortality risk among patients with CHF. Importantly, a U-shaped relation between RPR and in-hospital mortality emerged. Subsequent prospective investigations or randomized controlled trials are requisite to consolidate causal inference.

## Data Availability

All relevant data generated and analyzed during this study are available from the corresponding author upon reasonable request.

## Data availability

Publicly available datasets were analyzed in this study. This data can be found here: https://doi.org/10.5061/dryad.tx95x6b18 and https://doi.org/10.1136/bmjopen-2021059761.

---

**Figure S1.**
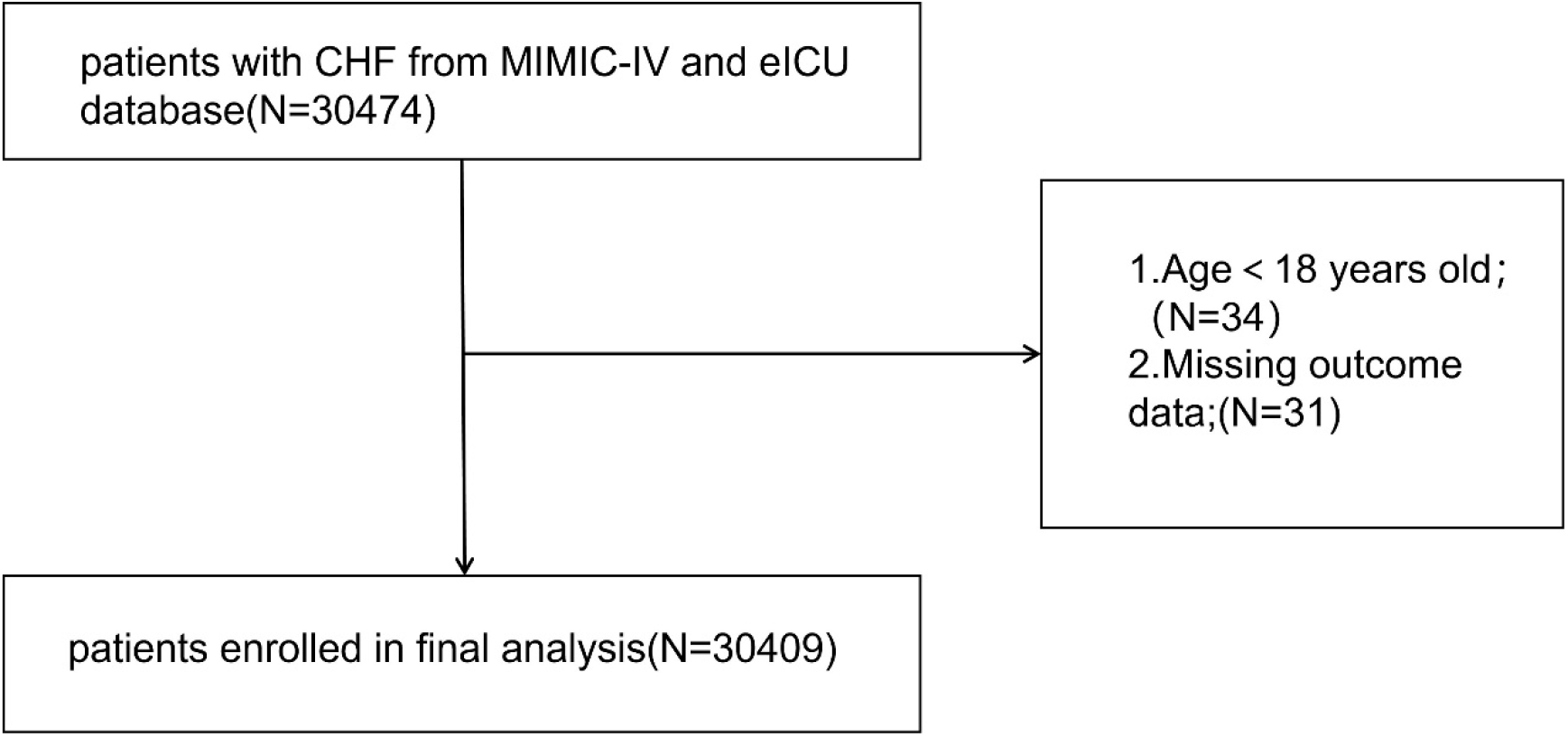
(Supplementary Materials Figure 1): Flow chart of the study population inclusion.

